# Mental illness stigma in England: What happened after the Time to Change Programme to reduce stigma and discrimination?

**DOI:** 10.1101/2024.02.20.24303075

**Authors:** Amy Ronaldson, Claire Henderson

## Abstract

**Background:** We investigated the extent to which positive changes in stigma outcomes reported over the course of Time to Change were sustained by 2023, two years after the programme’s end in 2021.

**Methods:** We used regression analyses to evaluate trends in outcomes. Measures were of stigma-related knowledge (Mental Health Knowledge Schedule (MAKS)), attitudes (Community Attitudes to the Mentally Ill scale (CAMI)), and desire for social distance (Reported and Intended Behaviour Scale (RIBS)). We also examined willingness to interact with people based on vignettes of depression and schizophrenia, and attitudes towards workplace discrimination against people with these conditions, using data from the British Social Attitudes Survey (BSAS) 2015 for comparison.

**Findings:** Reported in standard deviation units (95% confidence intervals (CI)), attitudes towards mental illness improved between 2008 and 2023 (SD=0.24, 95% CI=0.16 to 0.31), but following an increase of 9.9% between 2008-19, scores decreased by 3.3% (p=0.015). After improvements to 2019, 2023 MAKS and RIBS scores no longer differed from 2009 scores, indicating decreases since 2019 in stigma-related knowledge (MAKS scores declined 7.8% since 2019, p<0.001) and willingness to interact (RIBS scores declined by 10.2% since 2019, p<0.001). Conversely, comparison with BSAS 2015 data indicated that in 2023 respondents were more willing to interact with people with depression (β=-2.69, p<0.001) and schizophrenia (β=-2.70, p<0.001); and more likely to agree that people with either condition are just as likely to be promoted, and to disagree that their medical history should influence this. This change was most pronounced for schizophrenia (OR=2.52, 95% CI=2.02 to 3.14).

**Conclusions:** The lasting positive changes reflect support for non-discrimination and willingness to interact with someone after a sense of familiarity is evoked. Besides the end of Time to Change, interpretations for declines in other outcomes include the impacts of the covid-19 pandemic; economic stress; and reduced access to healthcare.

## Introduction

Stigma and discrimination against people with mental illness have substantial public health impact, contributing to inequalities^1^ including: poor access to mental and physical healthcare^2^; reduced life expectancy^3^; exclusion from higher education and employment^4^; increased contact with criminal justice systems; victimisation^5^; poverty and homelessness. There is growing investment in and evidence for the effectiveness of anti-stigma interventions, including programmes targeted at the general population and/or specific groups^6^.

In England, a programme against stigma and discrimination, Time to Change^7^, was delivered by the charities Mind and Rethink Mental Illness between 2007-21. Its first two phases ran from 2007-2011 and 2011-15, including a social marketing campaign launched in January 2009 aimed at adults aged 25-44 in middle income groups, and work with target groups. To evaluate Time to Change’s effect on public stigma, in 2009 measures of stigma related knowledge and desire for social distance were added to the pre-existing national Attitudes to Mental Illness survey^8^.

Between 2009-2015, there were significant improvements in stigma-related knowledge, attitudes, desire for social distance^9^. Surveys of mental health service users showed evidence for a reduction between 2008-14 in direct experiences of discrimination across multiple areas of life, particularly informal relationships^10^.

Time to Change phase 3 ran 2016-2021. The social marketing campaign from 2017 was aimed again at those aged 25-44, in an income group overlapping but lower than before and focussed on men’s mental health. The campaign promoted empathy as a key mediator of the effect of contact on prejudice^11^ while encouraging people to maintain contact^12^ (as opposed to avoidance). In the process, the campaign delivered parasocial (virtual) contact^12^ and promoted imagined contact^13^.

We previously reported positive change between 2008-19 in stigma-related knowledge, attitudes to mental illness, and desire for social distance from people with mental illness among the adult general population of England, supporting the effectiveness of the social marketing campaign^14^.

Other research conducted during the COVID-19 pandemic^15^ found an increase in desire for social distance from people with mental illness between March 2020 and March 2022, together with a decline in other measures designed to capture aspects of mental health literacy^16^.

We therefore investigated the extent to which the changes reported from 2008 to 2019 were sustained by 2023, two years after the end of Time to Change in England. We also wished to address a limitation of the measures, which enquire about mental illness or mental health problems in general, since other research has shown different changes over time in desire for social distance for depression versus schizophrenia^17^, and different patterns of change in newspaper coverage between schizophrenia and other disorders^18^. Therefore we also compared attitudes and desire for social distance towards people with depression and schizophrenia in 2023 with those available from one time point during Time to Change, by repeating measures used only in the 2015 British Social Attitudes Survey^19^.

## Methods

### Data source

The Attitudes to Mental Illness survey (AMI) was carried out annually in England from 2008 to 2017 and every 2 years since 2017 by Kantar TNS, and previously intermittently from 1994. Approximately 1700 respondents take part. A quota sampling frame is used to ensure a nationally representative sample of adults (16 years or older) living in England, and respondents are not resampled in later surveys. Detailed information about sampling methods^20^ can be obtained via the authors. Until 2019 respondents were interviewed face-to-face at home by trained personnel. Since then, data have been collected using address-based online surveying (https://www.kantar.com/-/media/Project/Kantar/Global/Expertise/Policy-and-Society/Address-Based-Online-Surveying.PDF) which offers web or paper self-completion. Measures of stigma-related knowledge and desire for social distance were added in 2009, just before the Time to Change social marketing campaign launched; therefore, the baseline for attitudes is 2008 and for the other outcomes is 2009.

### Measures

#### Stigma-related knowledge

Stigma-related knowledge was measured using the first six items of the Mental Health Knowledge Schedule (MAKS)^21^. covering: help seeking, recognition, support, employment, treatment and recovery. The standardised total score was used. These items were rated on a scale ranging from 1 (strong disagreement) to 5 (strong agreement) with higher scores indicating greater knowledge. The Cronbach’s alpha for the total scale in the current study was 0.59. This relatively low internal consistency has been reported previously^14^ and likely reflects the different knowledge domains measured by each item^21^.

#### Attitudes to mental illness

Attitudes to mental illness were measured using 26 of the 40 items of the Community Attitudes towards the Mentally Ill scale (CAMI)^22^ plus an item on employment-related attitudes added when the UK Department of Health first commissioned the survey in 1994^8^. The CAMI has factors: ‘Prejudice and Exclusion’; and ‘Tolerance and Support for Community Care’^23^. All items were rated on a scale ranging from 1 (strong disagreement) to 5 (strong agreement). The standardised scores of the total CAMI, ‘Prejudice and Exclusion’ and ‘Tolerance and Support for Community Care’ subscales were used. Higher scores indicate less stigmatising attitudes to mental illness. The internal consistency of the CAMI in this dataset was 0.87 (Cronbach’s alpha) for the total scale, and 0.85 and 0.76 for ‘Prejudice and Exclusion’ and ‘Tolerance and Support for Community Care’ respectively.

#### Desire for social distance

Desire for social distance was measured using the four items of the Reported and Intended Behaviour Scale (RIBS)^24^ which constitute the Intended Behaviour subscale and assess desire for social distance in terms of living with, working with, living nearby, and continuing a relationship with someone with a mental illness. These items were rated on a scale ranging from 1 (strong disagreement) to 5 (strong agreement) with higher scores indicating less desire for social distance. The total score was standardised. The internal consistency of the four items was 0.84 (Cronbach’s alpha).

#### Measures from the British Social Attitudes Survey 2015

For the first time in the Attitudes to Mental Illness survey, we included items used in the 2015 British Social Attitudes Survey (BSAS) conducted by the National Centre for Social Research (https://natcen.ac.uk/british-social-attitudes). This annual survey is designed to produce a representative sample of adults aged 18 or over living in private households. These data were accessed directly from the UK Data Service (https://beta.ukdataservice.ac.uk/datacatalogue/series/series?id=200006). We included BSAS participants living in England who responded to vignettes of a common mental health problem (depression) and a less common problem (schizophrenia). This allowed us to compare desire for social distance between 2015 and 2023. Two different people were described, and respondents were asked how willing they would be to interact with them in a range of situations (e.g., spend time socialising, move next door to). The vignettes were not labelled ‘depression’ or ‘schizophrenia’. We also repeated questions assessing expectations of and attitudes towards workplace discrimination against people with depression and schizophrenia as compared to a physical health comparator (diabetes). Specifically, respondents were asked about the likelihood of people with these conditions to be promoted, who had had repeated periods of time off work but whose illness was now under control through medication and were asked whether medical history should make a difference.

#### Familiarity with someone with a mental health problem

Familiarity with someone with a mental health problem was measured with the item: *Who is the person closest to you who has or has had some kind of mental illness*?’ Responses included self, immediate family, partner, other family, friend, acquaintance, work colleague, other, or no-one. The response to this item was categorised into three groups: self, other, none.

#### Sociodemographic variables

Several sociodemographic variables were included in the analysis: age, gender, ethnicity (Asian, Black, Other, White), socioeconomic position, and government office region. Socioeconomic position was based on the chief income earner of each household using the Market Research Society’s classification system (AB, C1, C2, DE). AB represents professional /managerial occupations, C1 represents other non-manual occupations, C2 represents skilled manual occupations, and DE represents semi-/unskilled manual occupations and people dependent on state benefits. Government office region is the lowest level information on participants’ location as described by the UK Government’s Office for National Statistics (ONS) (see Table 1).

**Table 1.** Participant demographics by survey year, un-weighted frequency and weighted per cent.

|  | 2008<br>(n=1703) | 2009<br>(n=1751) | 2010<br>(n=1745) | 2011<br>(n=1741) | 2012<br>(n=1717) | 2013<br>(n=1727) | 2014<br>(n=1714) | 2015<br>(n=1736) | 2016<br>(n=1765) | 2017<br>(n=1720) | 2019<br>(n=1785) | 2021<br>(n=1471) | 2023<br>(n=1638) |
| --- | --- | --- | --- | --- | --- | --- | --- | --- | --- | --- | --- | --- | --- |
| Gender, n (%) |  |  |  |  |  |  |  |  |  |  |  |  |  |
| *Female* | 925 (51.7) | 939 (51.5) | 939 (51.7) | 912 (51.5) | 924 (51.3) | 926 (51.0) | 893 (50.9) | 919 (51.6) | 918 (51.4) | 938 (50.8) | 933 (51.1) | 877 (50.9) | 1002 (51.3) |
| *Male* | 778 (48.3) | 812 (48.5) | 806 (48.3) | 829 (48.5) | 793 (48.7) | 801 (49.0) | 821 (49.1) | 817 (48.4) | 847 (48.6) | 782 (49.2) | 852 (48.9) | 570 (49.1) | 606 (48.7) |
| Age, mean (SD) | 46.7 (18.9) | 46.0 (18.8) | 46.5 (18.4) | 46.4 (19.2) | 46.4 (19.1) | 45.9 (18.3) | 46.0 (18.8) | 46.4 (19.2) | 46.3 (19.7) | 43.7 (20.0) | 46.0 (20.4) | Not<br>available | Not<br>available |
| Age group, n (%) |  |  |  |  |  |  |  |  |  |  |  |  |  |
| *16-24* | 188 (13.2) | 247 (14.3) | 240 (14.6) | 235 (14.4) | 258 (14.6) | 289 (14.6) | 221 (14.4) | 242 (14.1) | 211 (13.6) | 220 (17.8) | 275 (14.5) | 215 (12.9) | 142 (12.8) |
| *25-44* | 562 (36.0) | 633 (35.9) | 540 (35.1) | 545 (35.4) | 580 (34.8) | 568 (36.1) | 514 (36.2) | 528 (35.3) | 597 (35.5) | 491 (37.4) | 521 (36.2) | 522 (33.2) | 496 (32.6) |
| *45-64* | 525 (32.1) | 512 (31.3) | 549 (31.5) | 499 (30.6) | 506 (31.3) | 486 (31.1) | 506 (30.6) | 488 (31.5) | 488 (31.7) | 507 (27.7) | 484 (30.9) | 402 (32.3) | 491 (31.8) |
| *65+* | 428 (18.7) | 359 (18.5) | 416 (19.4) | 462 (19.5) | 373 (19.3) | 384 (18.3) | 473 (18.7) | 478 (19.0) | 469 (19.3) | 502 (17.1) | 505 (18.5) | 302 (21.6) | 508 (22.8) |
| Ethnicity, n (%) |  |  |  |  |  |  |  |  |  |  |  |  |  |
| *Asian* | 90 (5.5) | 112 (6.2) | 136 (8.5) | 134 (8.1) | 160 (9.7) | 127 (7.9) | 105 (6.6) | 120 (6.7) | 121 (7.0) | 76 (5.5) | 97 (6.5) | 117 (8.4) | 93 (8.5) |
| *Black* | 73 (4.4) | 63 (3.4) | 88 (4.9) | 64 (3.8) | 67 (3.8) | 66 (3.7) | 69 (4.0) | 99 (5.3) | 83 (4.7) | 61 (4.5) | 82 (4.7) | 21 (1.4) | 29 (2.0) |
| *Other* | 28 (1.9) | 26 (1.4) | 18 (1.1) | 20 (1.1) | 31 (1.8) | 44 (2.6) | 26 (1.6) | 39 (2.3) | 42 (2.6) | 41 (3.1) | 57 (3.7) | 29 (1.9) | 54 (4.5) |
| *White* | 1503 (88.1) | 1542 (89.0) | 1496 (85.5) | 1504 (87.0) | 1449 (84.7) | 1474 (85.9) | 1507 (87.8) | 1472 (85.7) | 1507 (85.7) | 1529 (87.0) | 1535 (85.1) | 1281 (88.3) | 1423 (85.0) |
| Socio-economic<br>position, n (%) |  |  |  |  |  |  |  |  |  |  |  |  |  |
| *AB* | 315 (21.0) | 279 (19.4) | 300 (20.2) | 322 (20.5) | 292 (19.3) | 302 (20.5) | 353 (21.4) | 335 (22.2) | 271 (18.9) | 350 (22.4) | 291 (20.5) | 541 (39.8) | 593 (39.1) |
| *C1* | 433 (29.6) | 454 (32.2) | 464 (31.7) | 450 (29.8) | 456 (31.0) | 445 (30.4) | 457 (29.2) | 432 (28.4) | 430 (30.6) | 501 (35.3) | 443 (30.4) | 317 (21.8) | 308 (19.7) |
| *C2* | 363 (21.1) | 389 (20.8) | 342 (19.2) | 340 (21.1) | 368 (21.6) | 362 (20.8) | 333 (20.5) | 354 (20.4) | 371 (20.7) | 296 (15.4) | 383 (20.9) | 181 (12.1) | 195 (13.0) |
| *DE* | 592 (28.2) | 629 (27.6) | 639 (28.8) | 629 (28.6) | 601 (28.1) | 618 (29.1) | 571 (29.0) | 615 (29.1) | 693 (29.8) | 573 (26.9) | 668 (28.3) | 423 (26.4) | 520 (28.2) |
| Familiarity with<br>mental health<br>problems, n (%) |  |  |  |  |  |  |  |  |  |  |  |  |  |
| *Self* | 102 (6.0) | 92 (5.0) | 75 (4.2) | 90 (5.6) | 111 (6.4) | 120 (6.6) | 126 (7.4) | 124 (6.9) | 124 (7.4) | 143 (9.2) | 152 (8.9) | 253 (17.5) | 229 (15.4) |
| *Other* | 665 (42.5) | 902 (54.0) | 892 (53.0) | 896 (53.5) | 926 (55.9) | 963 (57.9) | 953 (57.5) | 963 (58.1) | 1013 (61.1) | 980 (58.8) | 929 (55.0) | 953 (70.4) | 1076 (69.9) |
| *None* | 846 (51.5) | 718 (41.0) | 738 (42.8) | 706 (41.0) | 645 (37.7) | 610 (35.5) | 606 (35.1) | 632 (35.0) | 586 (31.5) | 566 (32.0) | 659 (36.1) | 150 (12.2) | 232 (14.7) |
| Region, n(%) |  |  |  |  |  |  |  |  |  |  |  |  |  |
| *North East* | 86 (5.3) | 86 (5.0) | 88 (4.8) | 83 (4.8) | 95 (5.5) | 82 (4.9) | 76 (4.4) | 76 (5.5) | 98 (5.3) | 73 (4.6) | 86 (4.7) | 82 (4.8) | 104 (4.9) |
| *North West* | 218 (12.9) | 245 (14.2) | 244 (13.6) | 233 (13.6) | 235 (12.8) | 233 (13.6) | 240 (13.8) | 280 (20.5) | 242 (13.8) | 238 (13.6) | 250 (13.2) | 172 (12.9) | 248 (12.9) |
| *York and Hum* | 172 (9.8) | 174 (9.6) | 178 (9.9) | 176 (10.5) | 185 (10.3) | 174 (9.9) | 169 (10.6) | 131 (9.6) | 186 (10.3) | 142 (7.9) | 188 (10.7) | 156 (9.7) | 168 (9.7) |
| *East Midlands* | 157 (9.1) | 151 (8.7) | 147 (9.9) | 159 (8.5) | 129 (8.2) | 152 (8.6) | 140 (8.0) | 139 (7.2) | 143 (8.1) | 152 (7.8) | 154 (9.0) | 120 (8.6) | 148 (8.6) |
| *West Midlands* | 173 (9.7) | 192 (10.9) | 190 (10.5) | 195 (10.4) | 186 (10.9) | 188 (10.8) | 189 (10.6) | 167 (8.6) | 189 (10.6) | 188 (11.0) | 173 (9.7) | 145 (10.5) | 145 (10.4) |
| *East* | 186 (11.3) | 187 (10.4) | 193 (11.3) | 192 (10.6) | 178 (10.3) | 188 (11.0) | 202 (11.3) | 202 (10.5) | 191 (11.3) | 199 (10.8) | 196 (11.7) | 156 (11.1) | 204 (11.1) |
| *South East* | 287 (16.9) | 278 (16.7) | 282 (17.0) | 285 (16.9) | 291 (17.6) | 285 (16.0) | 287 (17.1) | 305 (16.0) | 290 (16.9) | 295 (17.1) | 283 (15.6) | 243 (16.3) | 263 (16.3) |
| *South West* | 161 (9.1) | 176 (10.1) | 173 (9.6) | 176 (10.5) | 170 (10.1) | 179 (10.4) | 164 (9.8) | 146 (7.3) | 176 (10.0) | 181 (10.5) | 171 (9.0) | 198 (10.1) | 215 (10.1) |
| *London* | 263 (16.0) | 262 (14.5) | 250 (14.8) | 248 (14.4) | 248 (14.4) | 246 (14.9) | 247 (14.6) | 290 (14.6) | 250 (13.8) | 252 (16.8) | 284 (16.4) | 199 (16.0) | 143 (16.0) |

### Statistical Analysis

Descriptive statistics for participant demographics and crude outcome scores were calculated and reported by survey year. All analyses were weighted by age, gender, and ethnicity to reflect population characteristics in England. Survey weights were taken from the ONS. For modelling, the quota sample was treated as a probability sample.

We used multiple regression models to evaluate patterns of change in (i) stigma related knowledge (MAKS scores), (ii) attitudes to mental illness (CAMI scores)and (iii) desire for social distance (RIBS Intended Behaviour). All models used the standardised scores of the measures as dependent variables meaning the outputs were interpreted in standard deviation units. All the models included a fixed effect for year using a categorical dummy variable. We used the distributional approach to obtain estimates for the proportion of the population whose outcomes changed between two comparative years (i.e. 2023 and baseline, 2023 and 2019)^25^. This method uses the parameters of the normal distribution and converts results from linear regression models into corresponding proportions, using the area under the standard normal curve. We included covariates to control for differences in sociodemographic factors: age group, gender, ethnicity, socioeconomic position, familiarity with someone with a mental health problem (self, other, none), government office region. The same analysis has been applied to previous AMI data ^9,14,26,27^, except that the government office region variable was not included during Phase 1 of Time to Change^27^. Interactions between year and sociodemographic factors (age group, gender, ethnicity, socioeconomic position, government office region) were tested to see whether patterns of change in outcomes over time differed by groups. The interaction terms were added separately to the initial models and evaluated for statistical significance using a Wald test.

To compare desire for social distance (vignettes) and attitudes towards workplace discrimination we merged and harmonised relevant data from BSAS 2015 with data from the Attitudes to Mental Illness survey 2023. Logistic regression models were used to assess change between 2015 and 2023in willingness to interact with a person with depression (‘Stephen’) or schizophrenia (‘Andy’). A binary outcome variable was created; ‘1’ indicated that someone was *fairly* or *very willing* and ‘0’ indicated *neither willing nor unwilling, fairly unwilling*, or *very unwilling*. Item responses were summed to generate overall scores, with lower scores indicating more willingness to interact with a person with depression or schizophrenia. Linear regression models were used to assess change over time in overall scores. Sociodemographic factors common to both datasets were included as covariates in the logistic regression models: age group, gender, ethnicity, socioeconomic position, and government office region.

Similarly, logistic regression models were used to assess changes in attitudes towards workplace discrimination between 2015 and 2023. A binary outcome was created relating to likelihood of promotion for each condition where ‘1’ indicated that a person would be just as likely and ‘0’ indicated that they would be *slightly or much less likely*. A binary outcome was also created relating to whether someone’s condition should make a difference to likelihood of promotion with ‘1’ indicating that they *definitely/probably should* and ‘0’ indicating that they *probably/definitely should not*.

All analyses were performed using STATA 17.0 (Stata Corp LLP, College Station, Texas, USA).

### Ethics

The King’s College London Psychiatry, Nursing and Midwifery Research Ethics Subcommittee exempted analysis of these survey data as secondary analysis of anonymised data.

## Results

### Sample characteristics

Participant demographics are reported by survey year in Table 1. Over time, the distribution of male and female participants and their regional distribution remains stable. Since the introduction of remote data collection (2021) there have been changes in some sociodemographic factors: more participants were 45 years or older, had professional/managerial occupations, and had experience with mental health problems (self and other). In 2021 and 2023, fewer Black respondents took part.

### Attitudes to mental illness

Figure 1 depicts the change over time in total CAMI scores by plotting the marginal estimates of the standardised scores by year. There have been improvements in total CAMI scores since 2008 (see Table 2). In 2023, participants scored 0.24 (0.16 to 0.31) SD units higher on the CAMI scale than in 2008 translating to a 9.4% improvement in attitudes (p<0.001). However, between 2019 and 2023 total CAMI scores declined by 3.3% (p=0.015). An interaction between year and both age and government region (adjusted Wald tests p<0.001) suggests that changes in CAMI scores over time differ according to these sociodemographic factors (Supplementary figure S1 and S2). Declines in scores since 2019 are most pronounced in those aged 45-64 years and in those residing in South East England.

**Table 2.** Multiple linear regression analyses of predictors of mental health-related attitudes, knowledge, and behaviour among the general public.

| Predictors | Attitudes: standardised CAMI scores (N=21,334) |  | Knowledge: standardised MAKs scores (N=19,726) |  | Intended behaviour (IB): standardised RIBS IB subscale scores (N=19,726) |  |
| --- | --- | --- | --- | --- | --- | --- |
|  | Standardised effect size (95% CI) | P value | Standardised effect size (95% CI) | P value | Standardised effect size (95% CI) | P value |
| Year |  |  |  |  |  |  |
| 2023 | 0.24 (0.16 to 0.31)* | <0.001 | 0.05 (-0.02 to 0.13) | 0.153 | 0.04 (-0.03 to 0.11) | 0.294 |
| 2021 | 0.32 (0.25 to 0.39)* | <0.001 | 0.11 (0.04 to 0.19)* | 0.003 | 0.14 (0.07 to 0.21)* | <0.001 |
| 2019 | 0.32 (0.26 to 0.38)* | <0.001 | 0.25 (0.18 to 0.32)* | <0.001 | 0.30 (0.23 to 0.36)* | <0.001 |
| 2017 | 0.24 (0.18 to 0.31)* | <0.001 | 0.16 (0.09 to 0.23)* | <0.001 | 0.29 (0.23 to 0.36)* | <0.001 |
| 2016 | 0.25 (0.18 to 0.32)* | <0.001 | 0.15 (0.09 to 0.22)* | <0.001 | 0.21 (0.14 to 0.27)* | <0.001 |
| 2015 | 0.20 (0.13 to 0.26)* | <0.001 | 0.16 (0.09 to 0.23)* | <0.001 | 0.17 (0.11 to 0.24)* | <0.001 |
| 2014 | 0.18 (0.11 to 0.24)* | <0.001 | 0.12 (0.05 to 0.19)* | <0.001 | 0.18 (0.12 to 0.25)* | <0.001 |
| 2013 | 0.08 (0.02 to 0.15)* | 0.013 | 0.03 (-0.04 to 0.09) | 0.418 | 0.11 (0.05 to 0.18)* | 0.001 |
| 2012 | 0.05 (-0.02 to 0.11) | 0.144 | 0.03 (-0.03 to 0.10) | 0.344 | 0.07 (0.01 to 0.14)* | 0.024 |
| 2011 | 0.01 (-0.05 to 0.08) | 0.656 | -0.01 (-0.08 to 0.05) | 0.700 | 0.03 (-0.04 to 0.09) | 0.392 |
| 2010 | 0.07 (0.00 to 0.13)* | 0.042 | -0.02 (-0.09 to 0.04) | 0.447 | 0.09 (0.03 to 0.16)* | 0.004 |
| 2009 (ref) | -0.00 (-0.07 to 0.06) | 0.925 | - | - | - | - |
| 2008 (ref CAMI) | - | - | - | - | - | - |
| Gender |  |  |  |  |  |  |
| Female | 0.17 (0.15 to 0.20)* | <0.001 | 0.16 (0.13 to 0.18)* | <0.001 | -0.00 (-0.03 to 0.02) | 0.870 |
| Male (ref) | - | - | - | - | - | - |
| Age |  |  |  |  |  |  |
| 16-24 | 0.03 (-0.01 to 0.07) | 0.206 | 0.03 (-0.01 to 0.08) | 0.179 | 0.53 (0.49 to 0.58)* | <0.001 |
| 25-44 | 0.12 (0.08 to 0.15)* | <0.001 | 0.15 (0.11 to 0.18)* | <0.001 | 0.45 (0.42 to 0.49)* | <0.001 |
| 45-64 | 0.21 (0.17 to 0.24)* | <0.001 | 0.22 (0.18 to 0.26)* | <0.001 | 0.40 (0.36 to 0.44)* | <0.001 |
| 65+ (ref) | - | - | - | - | - | - |
| Ethnicity |  |  |  |  |  |  |
| Asian | -0.42 (-0.47 to -0.37)* | <0.001 | -0.08 (-0.13 to -0.02)* | 0.005 | -0.40 (-0.47 to -0.34)* | <0.001 |
| Black | -0.35 (-0.41 to -0.28)* | <0.001 | -0.02 (-0.10 to 0.05) | 0.502 | -0.25 (-0.33 to -0.17)* | <0.001 |
| Other | -0.27 (-0.36 to -0.17)* | <0.001 | -0.07 (-0.17 to 0.03) | 0.196 | -0.28 (-0.38 to -0.17)* | <0.001 |
| White (ref) | - | - | - | - | - | - |
| Socioeconomic position |  |  |  |  |  |  |
| AB | 0.37 (0.33 to 0.40)* | <0.001 | 0.32 (0.28 to 0.36)* | <0.001 | 0.27 (0.23 to 0.31)* | <0.001 |
| C1 | 0.26 (0.23 to 0.30)* | <0.001 | 0.19 (0.15 to 0.23)* | <0.001 | 0.19 (0.16 to 0.23)* | <0.001 |
| C2 | 0.11 (0.08 to 0.15)* | <0.001 | 0.08 (0.04 to 0.12)* | <0.001 | 0.09 (0.06 to 0.13)* | <0.001 |
| DE (ref) | - | - | - | - | - | - |
| Familiarity with mental health |  |  |  |  |  |  |
| Self | 0.88 (0.83 to 0.93)* | <0.001 | 0.79 (0.74 to 0.85)* | <0.001 | 0.86 (0.81 to 0.90)* | <0.001 |
| Other | 0.55 (0.52 to 0.58)* | <0.001 | 0.45 (0.41 to 0.48)* | <0.001 | 0.56 (0.53 to 0.59)* | <0.001 |
| None (ref) | - | - | - | - | - | - |
| Region |  |  |  |  |  |  |
| North East | 0.28 (0.21 to 0.35)* | <0.001 | 0.09 (0.02 to 0.17)* | 0.015 | 0.18 (0.11 to 0.26)* | <0.001 |
| North West | 0.22 (0.17 to 0.27)* | <0.001 | 0.09 (0.03 to 0.14)* | 0.001 | 0.22 (0.17 to 0.27)* | <0.001 |
| York and Hum | 0.30 (0.24 to 0.35)* | <0.001 | 0.16 (0.10 to 0.22)* | <0.001 | 0.26 (0.20 to 0.32)* | <0.001 |
| East Midlands | 0.19 (0.13 to 0.24)* | <0.001 | 0.10 (0.04 to 0.17)* | 0.002 | 0.20 (0.14 to 0.25)* | <0.001 |
| West Midlands | 0.18 (0.13 to 0.23)* | <0.001 | 0.07 (0.02 to 0.13)* | 0.011 | 0.18 (0.13 to 0.24)* | <0.001 |
| East of England | 0.21 (0.16 to 0.26)* | <0.001 | 0.10 (0.04 to 0.15)* | 0.001 | 0.17 (0.12 to 0.23)* | <0.001 |
| South East | 0.16 (0.12 to 0.21)* | <0.001 | 0.16 (0.00 to 0.11)* | 0.033 | 0.12 (0.07 to 0.17)* | <0.001 |
| South West | 0.27 (0.22 to 0.32)* | <0.001 | 0.12 (0.06 to 0.18)* | <0.001 | 0.16 (0.11 to 0.22)* | <0.001 |
| London (ref) | - | - | - | - | - | - |

\*Significant at p<0.05 level

**Figure 1.**
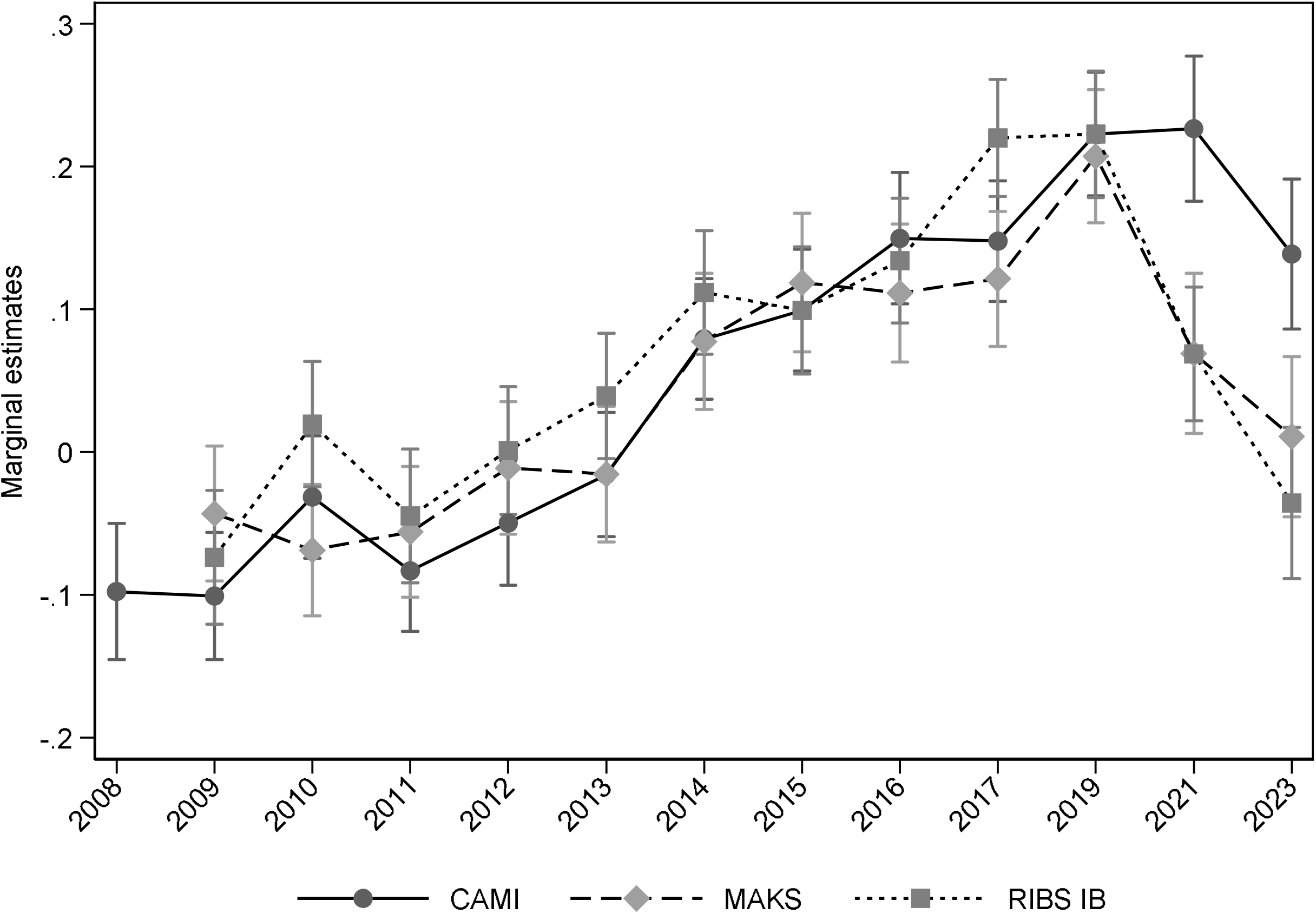
Marginal estimates of stigma-related attitudes (CAMI), knowledge (MAKS), and desire for social distance (RIBS intended behaviour) by year (95% CIs)

We also examined changes in CAMI ‘Prejudice and Exclusion’ and ‘Tolerance and Support for Community Care’ subscale scores (see Supplementary Table S1). Scores over time are presented in Supplementary figure S3. Scores on the ‘Prejudice and Exclusion’ subscale showed continued increases to 2021, but in 2023 scores returned to 2019 levels showing only a 0.1% improvement since that time (p=0.949). Since 2019, scores on the ‘Tolerance and Support for Community Care’ subscale decreased by 7.1% (p<0.001).

### Stigma-Related Knowledge

Changes in MAKS scores (marginal estimates of the standardised scores by year) are presented in Figure 1. Between 2009 and 2019, there were improvements in MAKS scores (see Table 2). Since 2019, MAKS scores have decreased by 7.8% (p<0.001) and results from 2023 no longer differ from 2009 scores (p=0.153). Interactions emerged between year and age (p=0.012), class (p=0.021), as well as region (p=0.010) (see Supplementary Figures S4 to S6). Specifically, since 2019 MAKS scores have declined in people aged under 25 years and aged 45 – 64 years, in people from the AB and C2 class groups, and in the North East, North West, South East and the South West of England.

### Desire for Social Distance

Figure 1 illustrates the change over time in total RIBS-IB scores (marginal estimates of the standardised scores). Significant improvements in scores were seen between 2009 and 2019 (Table 2). Since 2019, scores have decreased by 10.2% (p<0.001) and results from 2023 no longer differ from 2009 scores (p=0.294). Interactions emerged between year and socioeconomic position (p=0.017) and government region (p<0.001)(see Supplementary Figures S7 and S8). Declines in scores were seen in the AB and C2 socioeconomic group and in South East England since 2019.

### Comparison with British Social Attitudes Survey 2015

Sample characteristics for BSAS respondents in England are provided in Supplementary Table S2. Results from logistic regressions assessing changes in responses to individual items for each vignette (depression; schizophrenia) are presented in Supplementary Table S3. Linear regressions assessing change over time in overall vignette scores indicated that people in 2023 were more willing to interact with people with either depression (β=-2.71, p<0.001) or schizophrenia (β=-2.76, p<0.001) than in 2015.

Proportions of participant responses from the BSAS in 2015 and the AMI in 2023 are presented in Supplementary Table S4. Adjusted logistic regressions (Supplementary Table S5) revealed that, compared to people in 2015, people in 2023 were more likely to agree that people with depression and schizophrenia are just as likely to be promoted. This change was most pronounced for schizophrenia (OR=2.47, 95% CI=1.97 to 3.11). Moreover, people in 2023 were more likely to agree that having depression or schizophrenia should not make a difference to likelihood of promotion.

### Exploration of desire for social distance

To explore why we observed both decreases in RIBS-IB scores and greater willingness to have contact based on the vignettes representing individuals with symptoms of depression and schizophrenia, we graphically compared items from the RIBS-IB and the vignettes that tapped into similar aspects of life. As both the RIBS-IB and the vignettes were responded to on a 5-point Likert scale, we could calculate change scores by subtracting mean responses to each RIBS-IB and vignette item in 2015 from responses in 2023. These change scores are presented in Figure 2. Results indicate that RIBS-IB items concerning living with or near people with mental illness decreased or remained the same over this period, and items relating to working and being friends with people with mental illness increased marginally. Change scores for the vignettes suggest that there have been considerable increases in willingness to live, work, and be friends, particularly in relation to depression.

**Figure 2.**
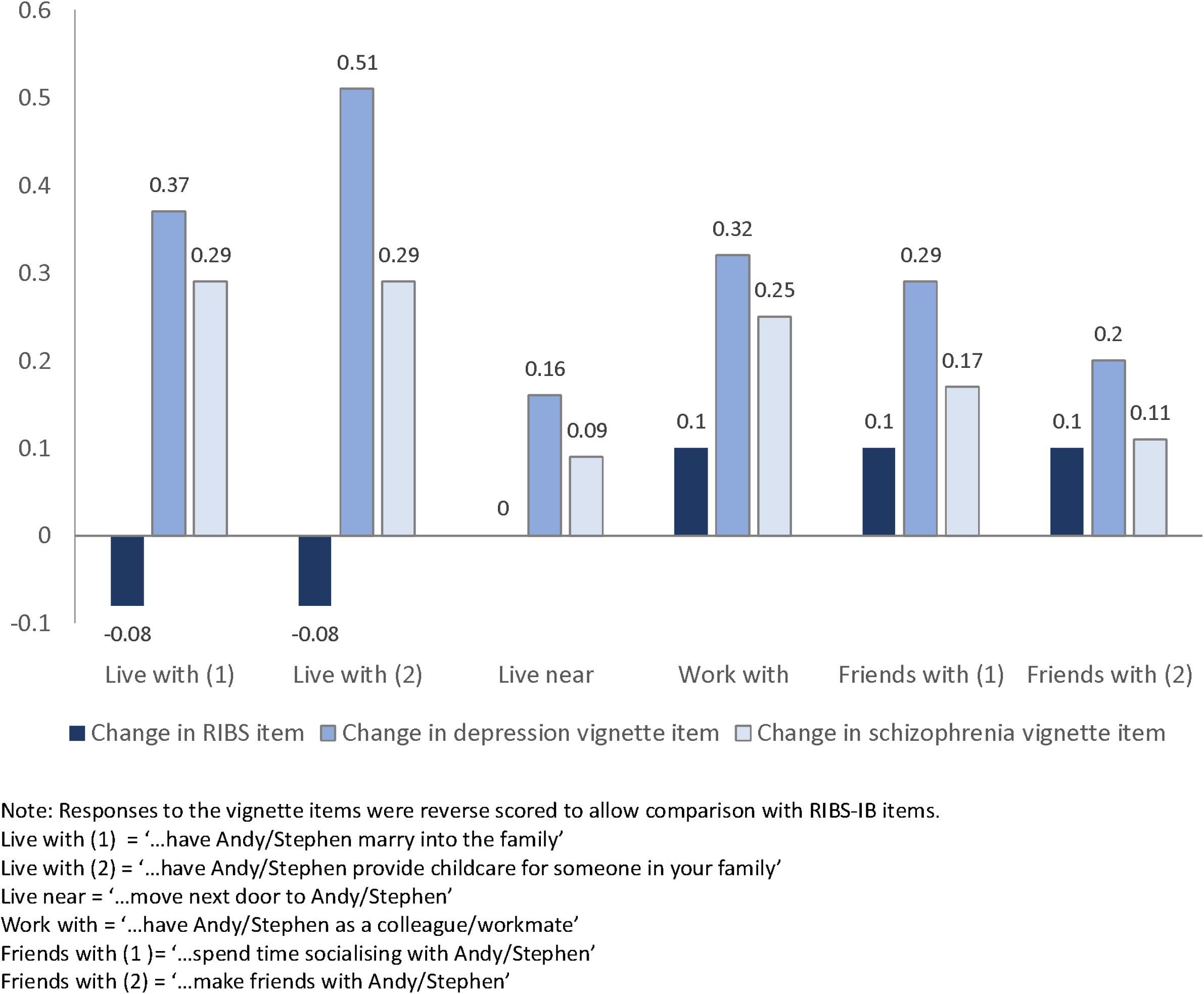
Change in RIBS-IB and vignette scores between 2015 and 2023

## Discussion

Our results show a complicated set of changes over time. Consistent with the surveys undertaken during the course of the Every Mind Matters campaign from 2019-22^15^, the stigma measures using questions about mental illness or mental health problems show an increase in stigma over this period following the improvements seen between 2008-19, such that although attitudes are still more positive since 2008, stigma related knowledge and desire for social distance began to decline before the end of Time to Change and are the same as in 2009. In contrast, the questions based on vignettes of men with depression or schizophrenia and those regarding workplace treatment showed reduced desire for social distance and more support for and expectation of fair workplace treatment, for someone with experience of depression and schizophrenia since 2015. While a greater proportion of people think that medical history should make a difference in relation to promotion for someone with schizophrenia compared to depression and diabetes, views regarding each condition have converged.

Consistent with this, the measure of stigma in relation to mental illness in general showing least deterioration is the Prejudice and Exclusion subscale of the CAMI, which also reflects awareness of rights and values of non-discrimination. In contrast, the reduction in the tolerance and support for community care subscale may reflect reduced support for government spending on mental health care at a time when personal taxation as a percentage of national income is at its highest since 1948.

The decline in stigma-related knowledge since 2019 reveals more therapeutic pessimism and less confidence in being able to help. This may reflect the greater difficulties in access to mental health care during and since the pandemic, as well as in self-management of mental health due to reduced access to salutogenic processes during lockdowns, and subsequently due to inflation.

The contrast between the negative changes in the RIBS-IB and the positive changes in response to the vignette questions is harder to explain, as the questions are similar. One interpretation is that the AMI survey sample report generally more positive attitudes to 2019 than those covered by the BSAS, due to different sampling and/or data collection methods. However, both the BSAS 2015 and AMI survey to 2019 were conducted face to face, a method which may result in more socially desirable responses^28^. Hence, while the more negative attitudes found in 2021 and 2023 as compared to previous AMI results could be interpreted as due to this change in methods, this does not explain the positive change seen between face-to-face responses to the depression and schizophrenia vignettes in 2015 and online responses to the same questions in 2023. Further, the increase in desire for social distance measures using RIBS-IB after March 2020 during the evaluation of Every Mind Matters was observed using an online survey throughout.

We should therefore consider whether there has been a change in how people respond to vignettes about individuals such that less desire for social distance is expressed, while the RIBS-IB questions about mental illness in general among groups of people elicit increasingly negative responses since 2019. The former may reflect a more lasting impact of Time to Change, which promoted supportive contact with family, friends and colleagues experiencing a mental health problem^29^; vignettes about an individual using their first name create a sense of familiarity. The latter may reflect a greater desire to avoid others who are unknown. While fear of contagion may have influenced responses in 2021, this seems less likely for 2023. However, responses to the RIBS-IB might be more affected than the vignette questions and for longer by personality changes identified later during the pandemic among adults. These include reductions in agreeableness and conscientiousness, although these data are from the USA^30^.

### Strengths and limitations

We calculated sampling errors even though a quota sample was used which violates some statistical assumptions but allowed us to calculate results as if the data were from a probability sample. While probability sampling has been used to measure single aspects of stigma in England at one time point ^19,23^ no current epidemiological survey has allowed repeated assessment of multiple aspects of stigma. Further, the lack of data for the BSAS items in 2019 means we cannot ascertain whether these attitudes have also worsened since 2019 after improving further after 2015. However, the analysis used nationally representative datasets and the demographic associations for attitudes are consistent with those found using the Health Survey for England 2014^23,31^. Although we included a significant number of confounders in our analyses, it is possible there were unmeasured confounders that may bias our results.

### Implications

Our results reinforce the multifaceted nature of mental illness-related stigma and the importance of measuring them, including in relation to common versus less common disorders, discrimination where legislation is relevant and intended interpersonal behaviour with versus without vignettes of named individuals. We predict that the aspects of stigma that worsened since 2019 will improve if and when economic conditions and access to treatment for common mental disorder improve. However, it remains to be seen whether the improvements seen over the course of Time to Change can be regained in the absence of such a programme.

## Data Availability

All data produced in the present study are available upon reasonable request to the authors

## Acknowledgements

We are grateful for collaboration on the evaluation by George Hoare and Alex Viccars at Mind.

