## Supplementary Material for "Mental illness stigma in England: What happened after the Time to Change Programme to reduce stigma and discrimination?"

| **Table S1.** Multiple linear regression analyses of predictors of CAMI ‘Prejudice/Exclusion’ and ‘Tolerance and Support for Community Care’ subscales | | | | |
| --- | --- | --- | --- | --- |
| Predictors | CAMI: Prejudice/Exclusion | | CAMI: Tolerance and support for community care | |
|  | Standardised effect size (95% CI) | P value | Standardised effect size (95% CI) | P value |
| Year |  |  |  |  |
| 2023 | 0.29 (0.23 to 0.36) | <0.001 | 0.10 (0.02 to 0.18) | 0.013 |
| 2021 | 0.35 (0.28 to 0.41) | <0.001 | 0.21 (0.14 to 0.29) | <0.001 |
| 2019 | 0.29 (0.23 to 0.35) | <0.001 | 0.28 (0.21 to 0.35) | <0.001 |
| 2017 | 0.22 (0.16 to 0.28) | <0.001 | 0.22 (0.15 to 0.29) | <0.001 |
| 2016 | 0.19 (0.12 to 0.25) | <0.001 | 0.27 (0.19 to 0.34) | <0.001 |
| 2015 | 0.13 (0.07 to 0.19) | <0.001 | 0.23 (0.16 to 0.30) | <0.001 |
| 2014 | 0.13 (0.07 to 0.20) | <0.001 | 0.19 (0.12 to 0.26) | <0.001 |
| 2013 | 0.03 (-0.03 to 0.09) | 0.402 | 0.13 (0.06 to 0.20) | <0.001 |
| 2012 | -0.02 (-0.08 to 0.04) | 0.531 | 0.13 (0.06 to 0.20) | <0.001 |
| 2011 | -0.06 (-0.012 to 0.01) | 0.070 | 0.11 (0.04 to 0.18) | 0.003 |
| 2010 | 0.01 (-0.05 to 0.07) | 0.776 | 0.13 (0.06 to 0.20) | <0.001 |
| 2009 | -0.07 (-0.14 to -0.01) | 0.021 | 0.09 (0.02 to 0.16) | 0.011 |
| 2008 (ref) | - | - | - | - |
| Gender |  |  |  |  |
| Female | 0.17 (0.15 to 0.20) | <0.001 | 0.12 (0.10 to 0.15) | <0.001 |
| Male (ref) | - | - | - | - |
| Age |  |  |  |  |
| 16-24 | 0.20 (0.16 to 0.24) | <0.001 | -0.21 (-0.26 to -0.17) | <0.001 |
| 25-44 | 0.24 (0.21 to 0.27) | <0.001 | -0.08 (-0.11 to -0.04) | <0.001 |
| 45-64 | 0.29 (0.25 to 0.32) | <0.001 | 0.05 (0.02 to 0.09) | 0.005 |
| 65+ (ref) | - | - | - | - |
| Ethnicity |  |  |  |  |
| Asian | -0.50 (-0.55 to -0.45) | <0.001 | -0.22 (-0.27 to -0.16) | <0.001 |
| Black | -0.37 (-0.43 to -0.30) | <0.001 | -0.23 (-0.31 to -0.16) | <0.001 |
| Other | -0.28 (-0.37 to -0.19) | <0.001 | -0.19 (-0.29 to -0.09) | <0.001 |
| White (ref) | - | - | - | - |
| Socioeconomic position |  |  |  |  |
| AB | 0.41 (0.37 to 0.44) | <0.001 | 0.22 (0.18 to 0.26) | <0.001 |
| C1 | 0.29 (0.26 to 0.33) | <0.001 | 0.16 (0.12 to 0.19) | <0.001 |
| C2 | 0.10 (0.07 to 0.14) | <0.001 | 0.10 (0.06 to 0.14) | <0.001 |
| DE (ref) | - | - | - | - |
| Familiarity with mental health |  |  |  |  |
| Self | 0.86 (0.81 to 0.91) | <0.001 | 0.69 (0.64 to 0.74) | <0.001 |
| Other | 0.56 (0.53 to 0.59) | <0.001 | 0.40 (0.37 to 0.43) | <0.001 |
| None (ref) | - | - | - | - |
| Region |  |  |  |  |
| North East | 0.26 (0.19 to 0.32) | <0.001 | 0.24 (0.16 to 0.31) | <0.001 |
| North West | 0.20 (0.15 to 0.25) | <0.001 | 0.19 (0.14 to 0.25) | <0.001 |
| York and Hum | 0.26 (0.20 to 0.31) | <0.001 | 0.28 (0.22 to 0.34) | <0.001 |
| East Midlands | 0.15 (0.10 to 0.21) | <0.001 | 0.19 (0.13 to 0.25) | <0.001 |
| West Midlands | 0.19 (0.14 to 0.24) | <0.001 | 0.12 (0.07 to 0.18) | <0.001 |
| East of England | 0.19 (0.14 to 0.24) | <0.001 | 0.18 (0.12 to 0.24) | <0.001 |
| South East | 0.16 (0.12 to 0.21) | <0.001 | 0.13 (0.07 to 0.18) | <0.001 |
| South West | 0.27 (0.22 to 0.32) | <0.001 | 0.20 (0.25 to 0.26) | <0.001 |
| London (ref) | - | - | - | - |

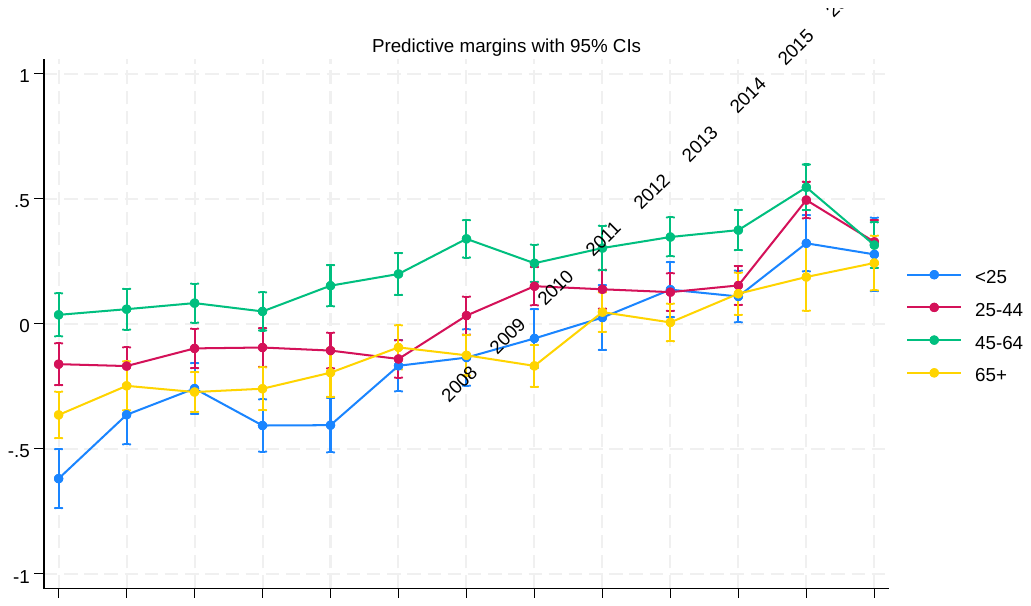

**Figure S1.** Marginal estimates of year*age group interaction (95% CIs) for mental health related attitudes (CAMI scores)

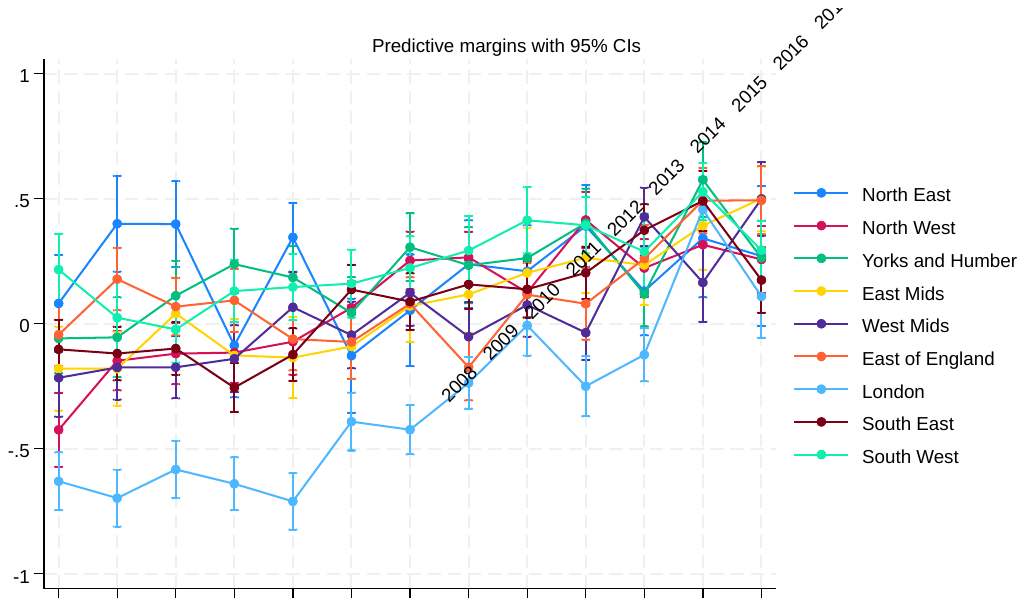

**Figure S2.** Marginal estimates of year*region interaction (95% CIs) for mental health related attitudes (CAMI scores)

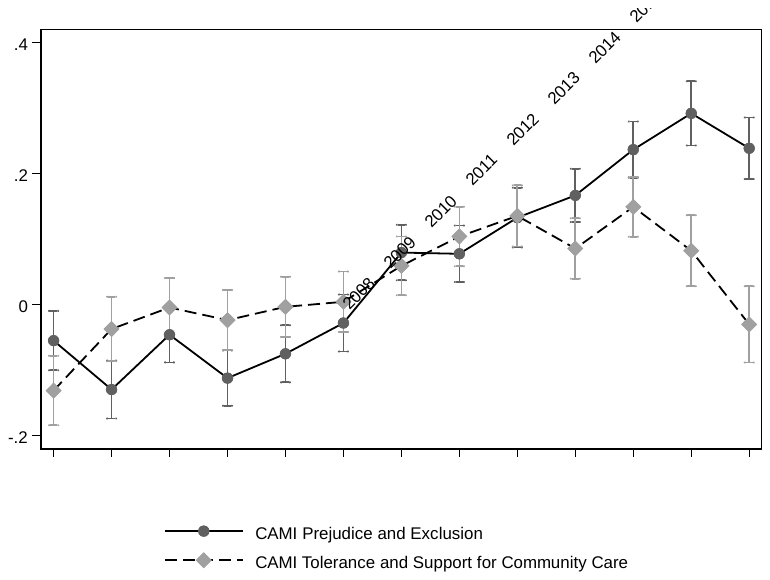

**Figure S3.** Marginal estimates of CAMI subscale scores by year (95% CIs)

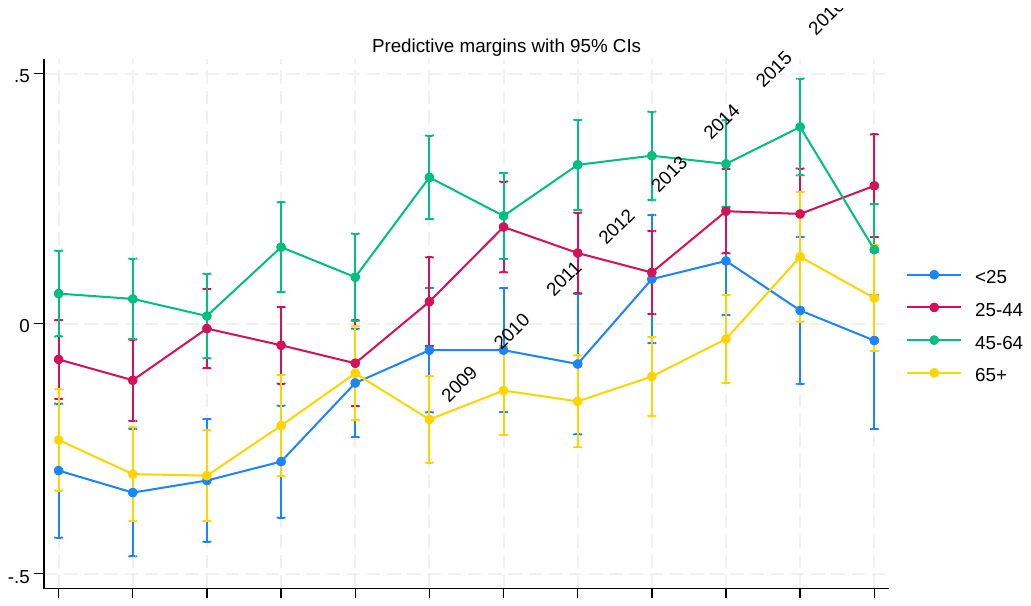

**Figure S4.** Marginal estimates of year*age group interaction (95% CIs) for mental health related knowledge (MAKS scores)

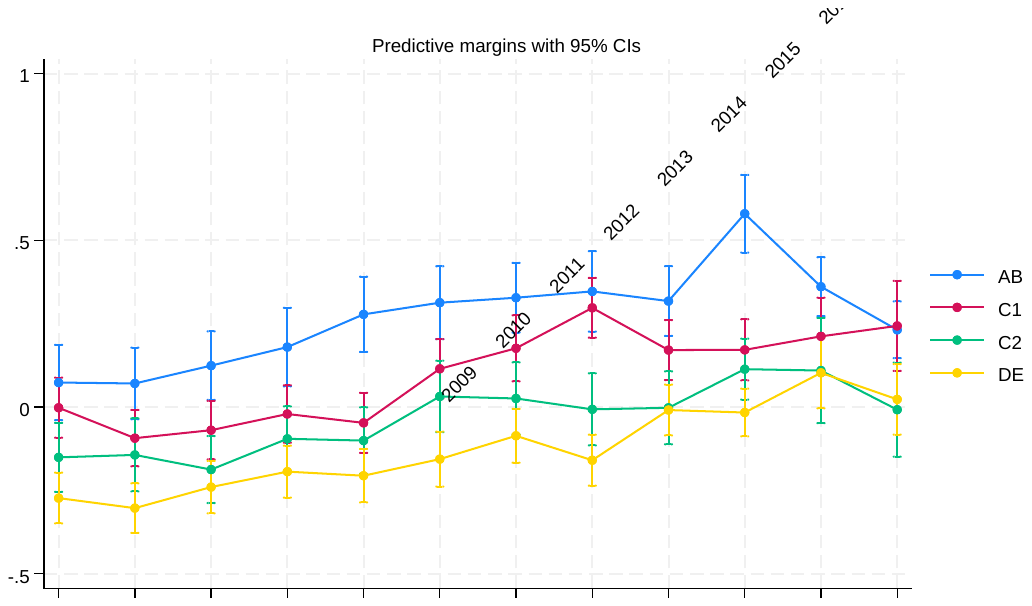

**Figure S5.** Marginal estimates of year*class interaction (95% CIs) for mental health related knowledge (MAKS scores)

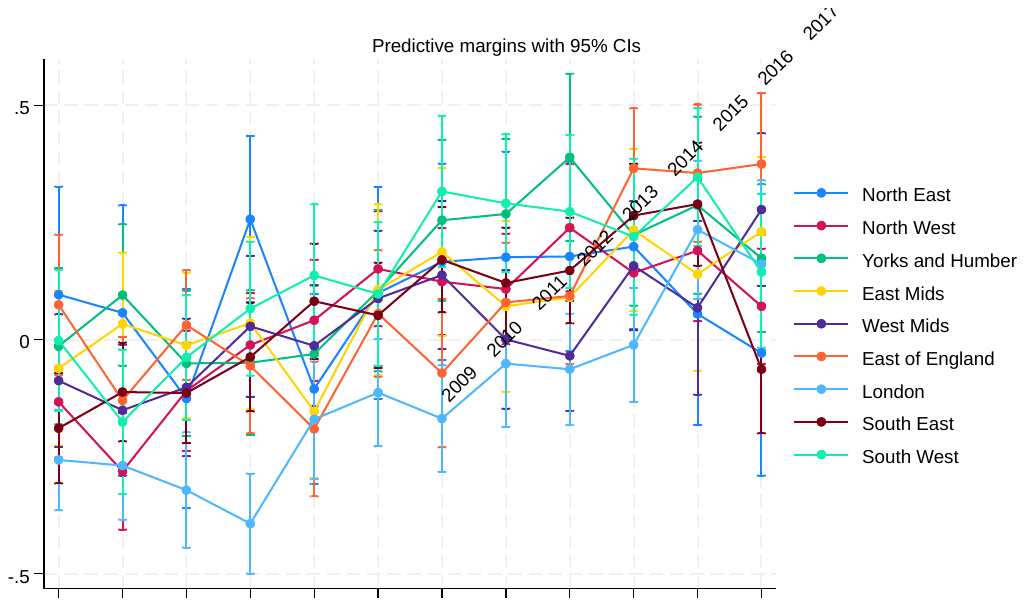

**Figure S6.** Marginal estimates of year*region interaction (95% CIs) for mental health related knowledge (MAKS scores)

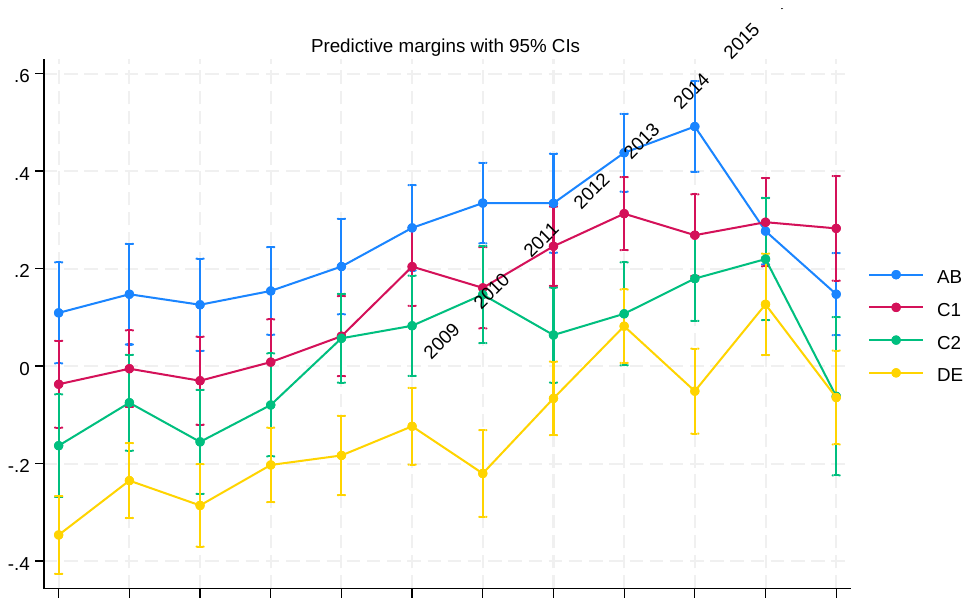

**Figure S7.** Marginal estimates of year*socioeconomic position interaction (95% CIs) for mental health related knowledge (RIBS IB scores)

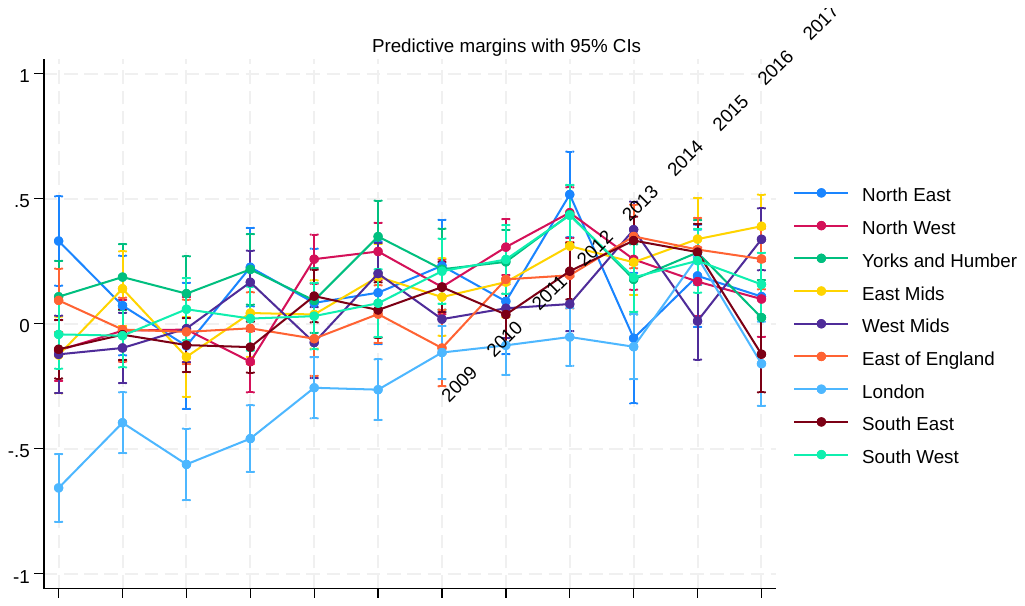

**Figure S8.** Marginal estimates of year*region interaction (95% CIs) for mental health related knowledge (RIBS IB scores)

| **Table S2.** BSAS 2015 sample characteristics for participants residing in England, N=1865 | |
| --- | --- |
|  | N(%) |
| Age group  Missing N=5 |  |
| *16-24y* | 131 (7.0) |
| *25-34y* | 279 (15.0) |
| *35-44y* | 334 (18.0) |
| *45-54y* | 337 (18.1) |
| *55-64y* | 294 (15.8) |
| *65+y* | 485 (26.1) |
| Gender  Missing N=0 |  |
| *Male* | 788 (42.2) |
| *Female* | 1077 (57.8) |
| Ethnicity  Missing N=2 |  |
| *Asian* | 111 (6.0) |
| *Black* | 68 (3.6) |
| *Other* | 34 (1.8) |
| *White* | 1650 (88.6) |
| Socioeconomic position  Missing N=92 |  |
| *AB* | 704 (39.70 |
| *C1* | 369 (20.8) |
| *C2* | 314 (17.7) |
| *DE* | 386 (21.8) |
| Local government region  Missing N=0 |  |
| *North East* | 103 (5.5) |
| *North West* | 263 (14.1) |
| *Yorks and Humber* | 172 (9.2) |
| *East Midlands* | 219 (11.7) |
| *West Midlands* | 200 (10.7) |
| *Eats of England* | 177 (9.5) |
| *London* | 207 (11.1) |
| *South East* | 349 (18.7) |
| *South West* | 175 (9.4) |

| **Table S3.** Logistic regressions assessing changes in willingness to interact with people with schizophrenia or depression between 2015 (BSAS) and 2023 (AMI) | | |
| --- | --- | --- |
| Vignette: Schizophrenia (Andy) | OR (95% CI) | p value |
| And now we would like you to think about how willing, or unwilling, you would be to ... |  |  |
| *…move next door to Andy?* | 1.01 (0.88 to 1.17) | 0.854 |
| *…spend time socializing with Andy?* | **1.22 (1.05 to 1.41)** | **0.007** |
| *…make friends with Andy?* | 1.07 (0.92 to 1.23) | 0.384 |
| *…have Andy as a colleague/workmate?* | **1.47 (1.27 to 1.70)** | **<0.001** |
| *…have Andy marry into the family?* | **1.66 (1.42 to 1.95)** | **<0.001** |
| *…have Andy provide childcare for someone in your family?* | **1.54 (1.23 to 1.93)** | **<0.001** |
| Vignette: Depression (Stephen) | OR (95% CI) | p value |
| And now we would like you to think about how willing, or unwilling, you would be to ... |  |  |
| *…move next door to Stephen?* | 1.13 (0.96 to 1.33) | 0.136 |
| *…spend time socializing with Stephen?* | **1.50 (1.28 to 1.76)** | **<0.001** |
| *…make friends with Stephen?* | **1.25 (1.06 to 1.47)** | **0.006** |
| *…have Stephen as a colleague/workmate?* | **1.60 (1.37 to 1.87)** | **<0.001** |
| *…have Stephen marry into the family?* | **1.68 (1.44 to 1.95)** | **<0.001** |
| *…have Stephen provide childcare for someone in your family?* | **2.29 (1.92 to 2.73)** | **<0.001** |
| Note: BSAS 2015 acts as reference category  AMI=Attitudes to Mental Illness; BSAS: British Social Attitudes Survey; CI=confidence interval; OR=odds ratio | | |

| **Table S4.** Changes in attitudes to mental health in the workplace measured in the British Social Attitudes Survey (BSAS 2015) and ‘Time to Change’ (2023) | | |
| --- | --- | --- |
|  | *BSAS 2015*  *N=2140* | *Time to Change 2023*  *N=1638* |
| *Views on promotion prospects* | 2015 | 2023 |
| **Depression** |  |  |
| % just as likely as anyone else to be promoted | 16.5 | 25.5 |
| % slightly less likely | 47.5 | 44.6 |
| % much less likely | 36.0 | 29.9 |
| % medical history definitely/probably should make a difference | 37.5 | 15.4 |
| **Schizophrenia** |  |  |
| % just as likely as anyone else to be promoted | 8.3 | 17.7 |
| % slightly less likely | 32.1 | 36.4 |
| % much less likely | 59.6 | 45.8 |
| % medical history definitely/probably should make a difference | 50.0 | 20.4 |
| **Diabetes** |  |  |
| % just as likely as anyone else to be promoted | 56.9 | 63.8 |
| % slightly less likely | 35.4 | 31.0 |
| % much less likely | 7.6 | 5.2 |
| % medical history definitely/probably should make a difference | 25.2 | 13.5 |

| **Table S5.** Logistic regressions assessing change in attitudes towards promotions in the workplace for people with depression, schizophrenia, or diabetes between 2015 (BSAS) and 2023 (AMI) | | | | | |
| --- | --- | --- | --- | --- | --- |
|  |  | Just as likely to be promoted | | Condition definitely/probably should not make a difference | |
|  |  | *OR (95% CI)* | *P value* | *OR (95% CI)* | *P value* |
| Depression | BSAS 2015 | Ref |  | Ref |  |
|  | AMI 2023 | 1.76 (1.49 to 2.11) | <0.001 | 1.48 (1.23 to 1.81) | <0.001 |
| Schizophrenia | BSAS 2015 | Ref |  | Ref |  |
|  | AMI 2023 | 2.47 (1.97 to 3.11) | <0.001 | 1.50 (1.25 to 1.81) | <0.001 |
| Diabetes | BSAS 2015 | Ref |  | Ref |  |
|  | AMI 2023 | 1.31 (1.13 to 1.52) | <0.001 | 1.38 (1.13 to 1.68) | 0.001 |
| AMI=Attitudes to Mental Illness; BSAS: British Social Attitudes Survey; CI=confidence interval; OR=odds ratio | | | | | |
